# From Genes to Personalized Cancer Care: A Systematic Review of Toxicity-Associated Genetic Variants in Solid Tumor Treatments

**DOI:** 10.1101/2025.10.21.25338446

**Authors:** Andrea Gonzalez-Hernandez, Alejandro Escamilla-Sanchez, Elisabeth Pérez-Ruiz, Alberto Rios, Cecilia A. Frecha, Felipe Vaca-Paniagua, Isabel Barragán, Sandra Perdomo, Antonio Rueda-Domínguez, Javier Oliver

## Abstract

**Background:** Pharmacogenomics has emerged as a crucial tool in precision medicine, offering the potential to personalise cancer treatments by predicting and managing therapy-induced toxicities. This systematic review examined the genetic basis of toxicities associated with radiotherapy, chemotherapy, and immunotherapy in solid tumours.

**Methods:** A comprehensive literature search was conducted across PubMed, Google Scholar, and PharmKB databases, covering the period from December 2019 to July 2024. This review focused on genetic variants linked to different treatment-related toxicities, including chemotherapy, radiotherapy, and immunotherapy, across various solid tumour types.

**Results:** The review primarily assessed immune-related adverse events and dermatologic, haematologic, neurological, and organ-specific toxicities (e.g. ototoxicity, hepatotoxicity, nephrotoxicity, and cardiotoxicity). This review highlights single-nucleotide variants (SNVs) as essential genetic markers for identifying treatment-related toxicities. However, data on many SNVs remains limited, highlighting the need for further research and clinical validation. These findings suggest that the understanding of genetic factors that contribute to toxicity may support treatment decisions, optimise patient outcomes, and promote advances in the field of precision oncology.

**Conclussion:** The identification of specific genetic variants could prevent the use of expensive and ineffective treatments and guide the selection of patients most likely to benefit from a specific therapy. Here, we provide valuable insights into the current state of knowledge regarding the genetic basis of toxicity in solid tumour treatments and emphasise the importance of integrating pharmacogenomics into personalised cancer care. To enhance patient outcomes and reduce the economic burden of cancer treatment, further research must validate these genetic markers and integrate the findings into clinical practice, thereby avoiding ineffective treatments for patients.

## Introduction

A key aspect of personalised precision medicine (PPM) is pharmacogenomics, which determines how genetic differences among individuals influence the safety and efficacy of drug treatments (1). Common genetic variants account for 20-30% of the differences in individual medical responses. They also play a crucial role in a patient’s susceptibility to treatment-related toxicity, influencing both the probability and severity of adverse events and the effectiveness of the treatment (2–4). These genetic differences can affect protein structure and lead to alternative drug metabolism, allowing for the prediction of drug response and toxicity. The treatment landscape of solid tumours has evolved rapidly through the integration of surgery, chemotherapy, radiotherapy (RT), targeted therapy, and immunotherapy. Recently, a large-scale cohort study of 75,000 individuals diagnosed with cancer found that 62.7% had pharmacogenetic variants in four genes (*DPYD*, *NUDT15*, *TPMT*, *and UGT1A1*) associated with adverse reactions to capecitabine, fluorouracil, mercaptopurine, thioguanine, and irinotecan, commonly used in cancer treatment (5).

Variations in specific genes can alter protein structures and drug metabolism pathways, allowing the prediction of treatment responses and identification of potential adverse events. As the landscape of solid tumour treatments has rapidly evolved to include surgery, chemotherapy, radiotherapy, targeted therapy, and immunotherapy, pharmacogenomics has become an essential tool for optimising these approaches. Pharmacogenomics helps minimise toxicity and improve therapeutic outcomes by tailoring treatments to the individual genetic profile.

Chemotherapy, the cornerstone of numerous common malignancies like breast, colorectal, and pulmonary neoplasms (6,7), often results in a range of adverse effects such as myelosuppression, nausea, vomiting, diarrhoea, hair loss, fatigue, and reproductive issues (6). Likewise, similar types of adverse events are observed with targeted therapy for the treatment of *HER2*-amplified breast carcinomas and *BRAF*-mutated melanomas; radiotherapy-treated patients suffering from solid tumors such as mammary, prostatic, pulmonary, or head and neck carcinomas may experience organ-specific toxicities according to the target organ, including mucositis, dysphagia, xerostomia (11,12), pneumonitis, dyspnea, or dry cough (13); and gastrointestinal toxicity, sexual dysfunction, and fertility concerns with pelvic radiation (14), in addition to psychological problems such as distress, anxiety, and depression (8–10). Finally, novel antitumour immunotherapeutic approaches designed to enhance the immune response against cancer cells, like immune checkpoint inhibitors (ICI), significantly improved the outcomes of melanoma, non-small cell lung cancer, and diverse solid tumours; however, but they can lead to a range of inflammatory side effects referred to as Immune-related Adverse Events (irAEs) that impede the continuation of treatment (15). The occurrence and nature of irAEs are multifactorial and include the type of ICIs used, tumour type, and clinical variables. However, the precise mechanisms underlying the occurrence of irAEs remain unclear.

As cancer treatment evolves, the ability to forecast and mitigate side effects has become increasingly vital, paving the way for more effective, patient-centred care in oncology. Early response monitoring, one one hand, may limit toxicity and on the other, enable the development of more precise treatment regimens (15,16), allowing healthcare providers to tailor treatments to individual patients, minimizing risks while maximizing therapeutic benefits. Additionally, it is crucial to consider the potential impact of the increasing number of cancer patients on the economic costs of cancer treatment and the management of health systems (17,18). Between 2010 and 2019, there was a significant increase in cancer burden, with 26% new cases, 20% deaths, and 16% increase in disability-adjusted life years (DALY) (19).

Identifying genetic variants that can guide personalised therapeutic approaches can help avoid the use of costly and ineffective treatments. The costs of these treatments vary significantly, with chemotherapy ranging from 125 to 150 USD per hour (20), radiotherapy (RT) costing between 7,000 and 13,000 USD (22,23), and ICI-based immunotherapy priced at approximately 8,900 USD per prescription (24). By tailoring therapies to individual genetic profiles, this approach facilitates a shift from generalised treatments to more predictable, effective, and cost-efficient interventions (25).

This systematic review thoroughly examined the literature on genetic variants that contribute to treatment toxicity in patients with solid tumours. The primary objective was to synthesise evidence and identify genetic markers associated with toxicity from chemotherapy, radiotherapy (RT), and immunotherapy, as well as to explore their implications for treatment outcomes. The findings aim to guide future research that could inform clinical decision-making and eventually support the integration of pharmacogenomics into personalised cancer care.

### Methodology

This review specifically targets original studies that investigated genetic variants associated with treatment-related toxicities in solid tumours, focusing on outcomes related to chemotherapy-, radiotherapy-, and immunotherapy-induced toxicities. A thorough literature review was conducted using reliable databases PubMed, Google Scholar, and PharmKB. Articles published between December 2019 and August 2024, specifically focusing on studies that investigated genetic markers associated with treatment toxicity in solid tumours, were included. Studies outside these treatment modalities, such as surgical outcomes, haematological malignancies, or non-genetic biomarkers, were excluded to maintain a focused examination of the pharmacogenomic landscape in solid tumour treatments.

No age restrictions were applied to the search, and all study designs were considered, excluding *in vitro* and *in vivo* studies in animal models. The search terms included “cancer,” “solid tumors,” “pharmacogenomics,” “toxicity markers,” “radiotherapy,” “chemotherapy,” “immunotherapy,” “irAEs,” “immune checkpoint inhibitors,” and “autoimmunity.” We examined all Single Nucleotide Variants (SNVs) across the entire human genome, including regulatory, exonic, and intronic regions. The credibility and conclusions of each article were assessed using the Critical Appraisal Skills Program (CASP) checklist (Oxford, UK, 2017). Data from each article were collected and summarised in an extraction form (Figure 1), including participants’ sociodemographic data, treatment intervention details, toxicology assessment, and statistical analysis outcomes of the examined genes.

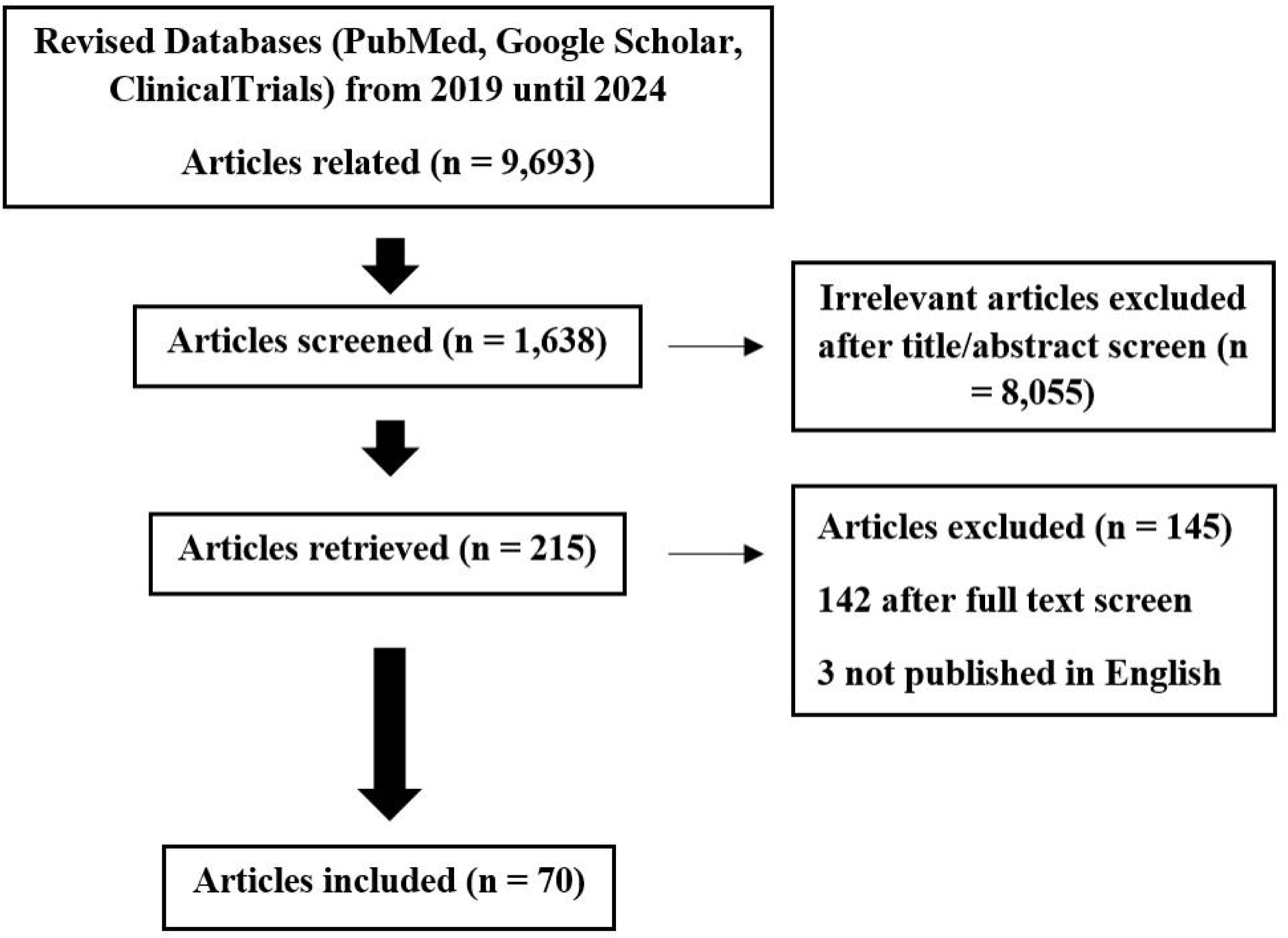

## Results

### Systematic review

The initial search yielded 9,693 original publications. After removing duplicated studies, 1,638 remained. Of these, 215 were selected for further review, and 70 studies were thoroughly analysed. The reviewed articles included 39 studies on chemotherapy, 12 on immunotherapy, 14 on radiotherapy, and 5 on chemoradiotherapy studies. Among these, 12 focused on irAEs, 19 on hematotoxicity, 12 on neurological toxicity, 7 on dermatologic toxicity, 18 on gastrointestinal toxicity, and 23 on other toxicities. In terms of methodology, most studies utilised PCR-based techniques, followed by microarrays and mass arrays; only nine studies used next-generation sequencing (NGS) approaches (Tables 1 and 2).

**Table 1.** Novel toxicological irAEs biomarkers recently discovered.

| Study | Cancer | Treatment | Toxicity associated | Associated genes | Variants | Methodology | N |
| --- | --- | --- | --- | --- | --- | --- | --- |
| Abdel-Wahab et al. 2021 (44) | Melanoma | ICI | irAEs | *ANKRD42*<br>*AGPS*<br>*CFAP65*<br>*GABRP*<br>*DSC2*<br>*BAZ2B*<br>*SEMA5A* | rs11743438<br>JHU_20.5718398<br>rs56328422<br>rs3026321 | Microarray | 89 |
| Ali et al. 2019 (38) | Metastatic Melanoma/Metastatic NSCLC | ICI | irAEs | *HLA* | DRB1*11:01<br>DQB1*03:01 | NGS | 102 |
| Ferguson et al. 2023 (40) | Melanoma | ICI | irAEs | *SYK* | rs7036417 | MassArray | 95D<br>97V |
| Groha et al. 2022 (43) | Multiple. Melanoma associated | ICI | irAEs | *IL7*<br>*IL22RA1*<br>*8q21 locus* | rs16906115<br>rs75824728<br>rs113861051 | GWAS | 1,751 |
| Iafolla et al. 2020 (37) | HNSCC, BC, Ovarian Cancer, Melanoma | ICI | irAEs | *HLA* | HLA-C | WES-NGS | 101 |
| Luo et al. 2021 (35) | NSCLC | ICI | irAEs<br>Immunotherapy-Mediated Thyroid Dysfunction | *HLA-DQA2* | rs9268543 | Microarray | 1,307 |
| Marschner et al. 2020 (46) | Various | ICI | irAes | *MIR146A* | rs2910164 | Taqman PCR | 179 |
| Montaudié et al. 2021 (39) | Metastatic Melanoma | ICI | irAEs | *ANO5*<br>*CFAP36*<br>*PNPT1*<br>*IL1RL1* | rs7481951<br>rs3762513<br>rs782637<br>rs78259<br>rs7882572<br>rs4988956 | Taqman PCR | 57D<br>57V |
| Refae et al. 2019 (41) | Advanced Cancer | ICI | irAEs | *UNG*<br>*IFNW1*<br>*CTLA4*,<br>*PD-L1*<br>*IFNL4* | rs246079<br>rs10964859<br>rs4143815<br>rs12979860<br>rs3087243<br>rs11571302<br>rs7565213 | MassArray | 94 |
| Taylor et al. 2022 (45) | Melanoma | ICI | irAEs | *IL7* | rs16906115 | GWAS | 214 |
| Udagawa et al. 2022 (42) | NSCLC<br>Melanoma,<br>HNSCC,<br>Gastric Cancer,<br>Kidney Cancer,<br>Hodgkin's<br>Lymphoma<br>Carcinoma of<br>Unknown<br>Primary,<br>Oesophageal<br>Cancer,<br>Rectal<br>Cancer,Mesothelioma,<br>Bone<br>Cancer,<br>Gastroesophageal<br>Junction<br>Cancer | ICI | irAEs | *21q21.3 near APP* | rs469490 | Microarray | 622 |
| Yano et al. 2020 (36) | Melanoma, NSCLC, Gastric Cancer | ICI | Pituitary irAEs | *HLA* | DR15<br>B52<br>Cw12 | Luminex | 11 |

**Table 2.**
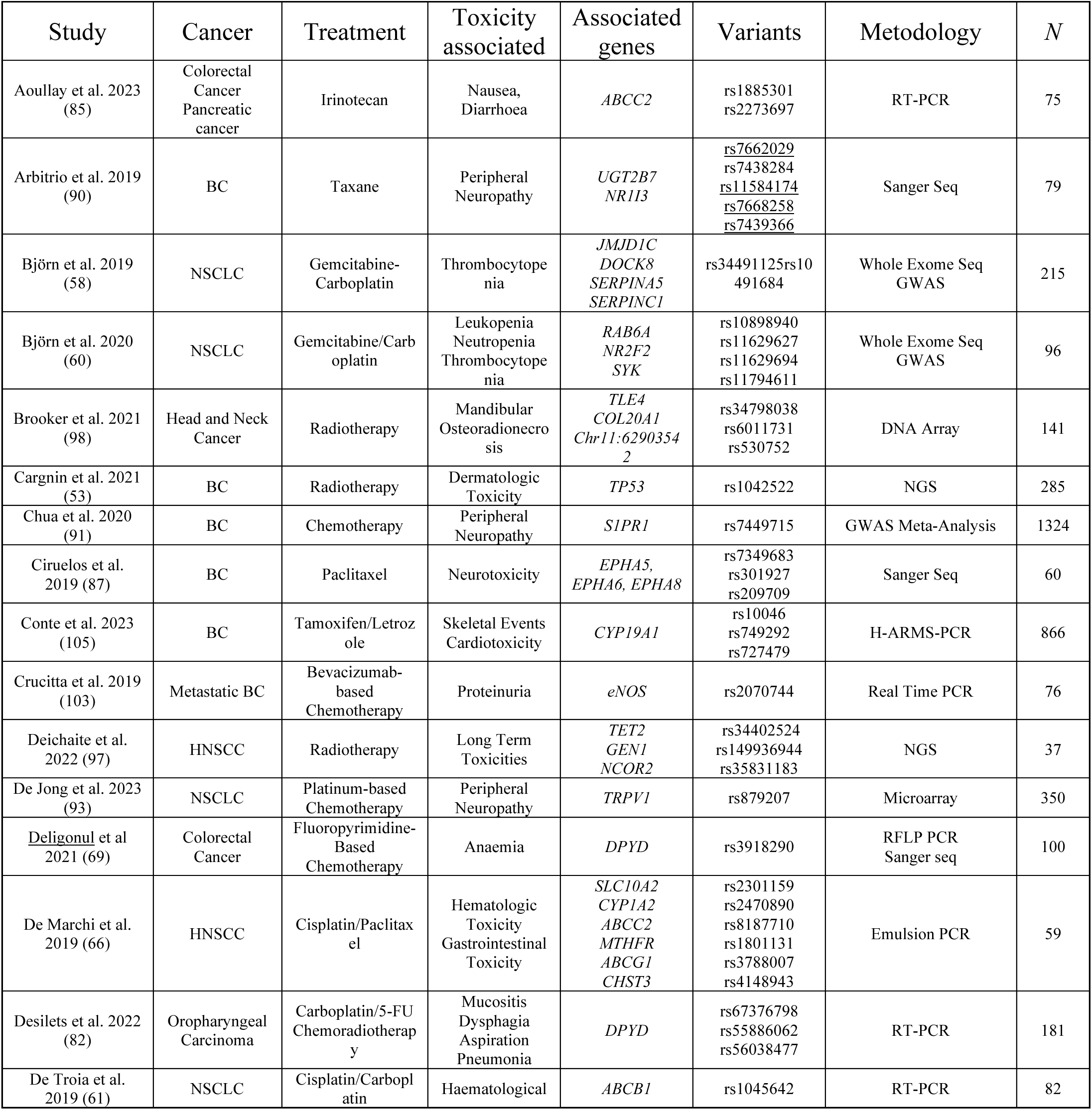
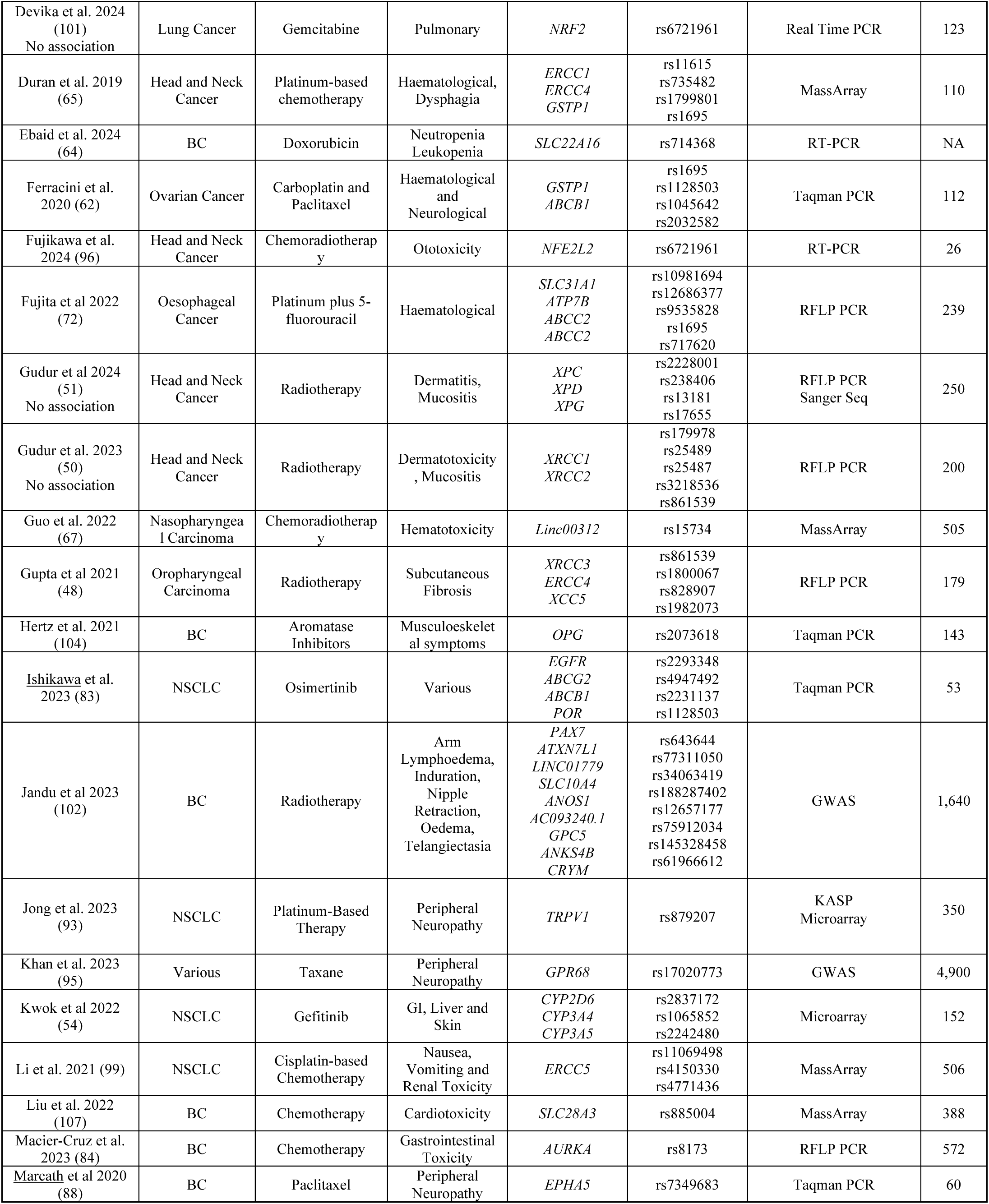
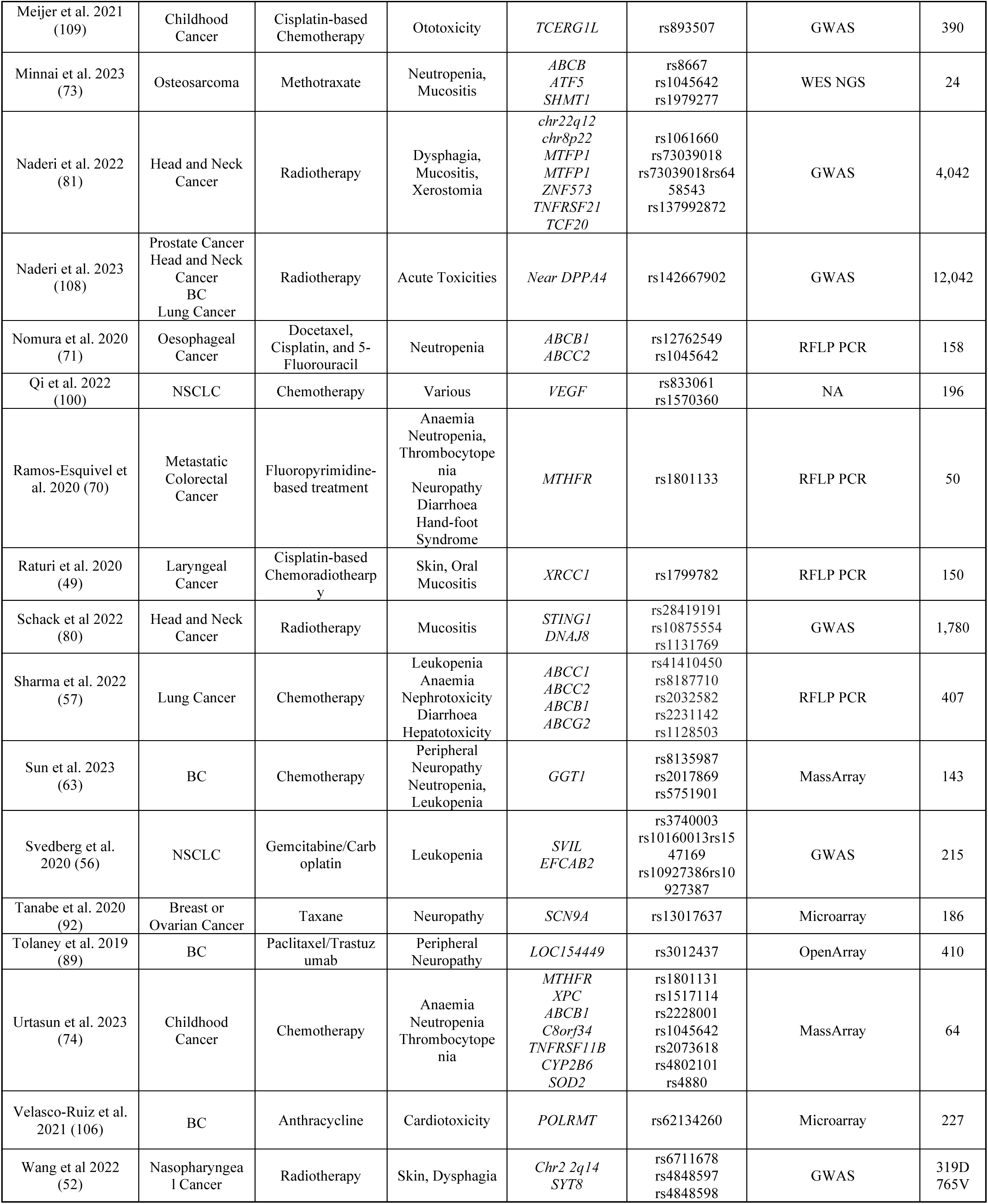
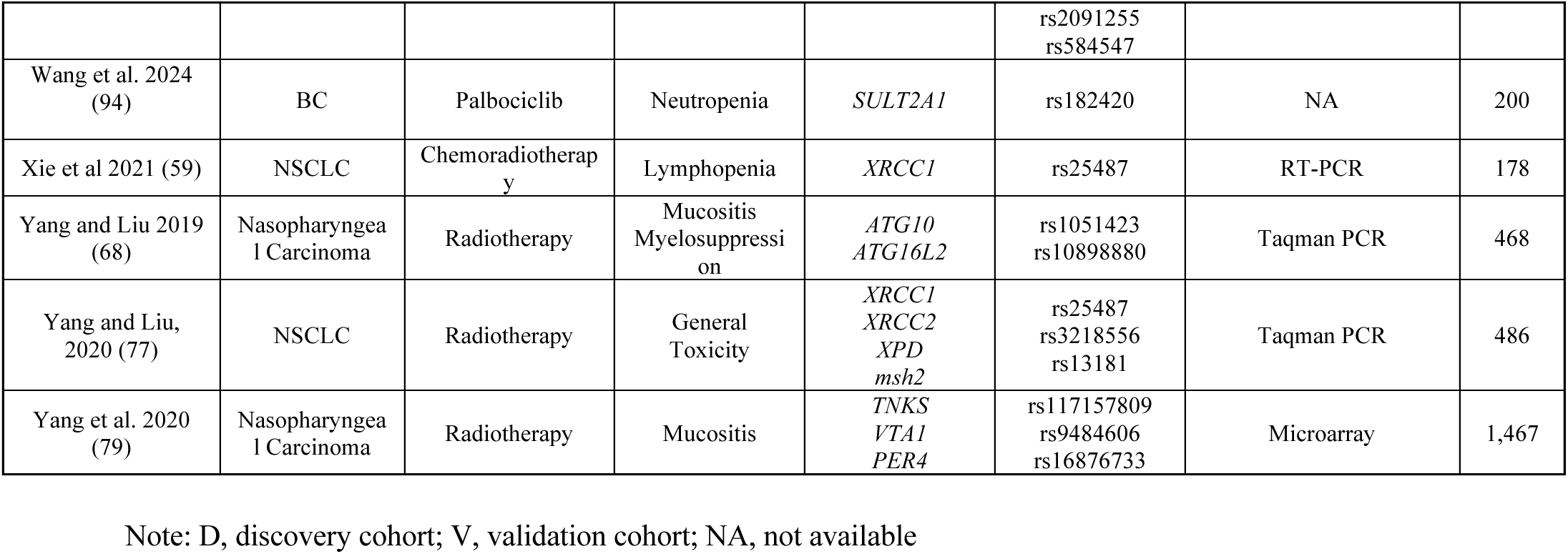
SNVs associated with toxicity to chemotherapy, radiotherapy, and targeted therapy.

Figure 2 represents a body map with the toxicity SNVs arranged by odds ratio (OR) and distinguishing risk (OR>6) from protective biomarkers (OR<0.3). Tables and a forest plot with more details are included in the Supplementary Material (Table S1 and Figure S1).

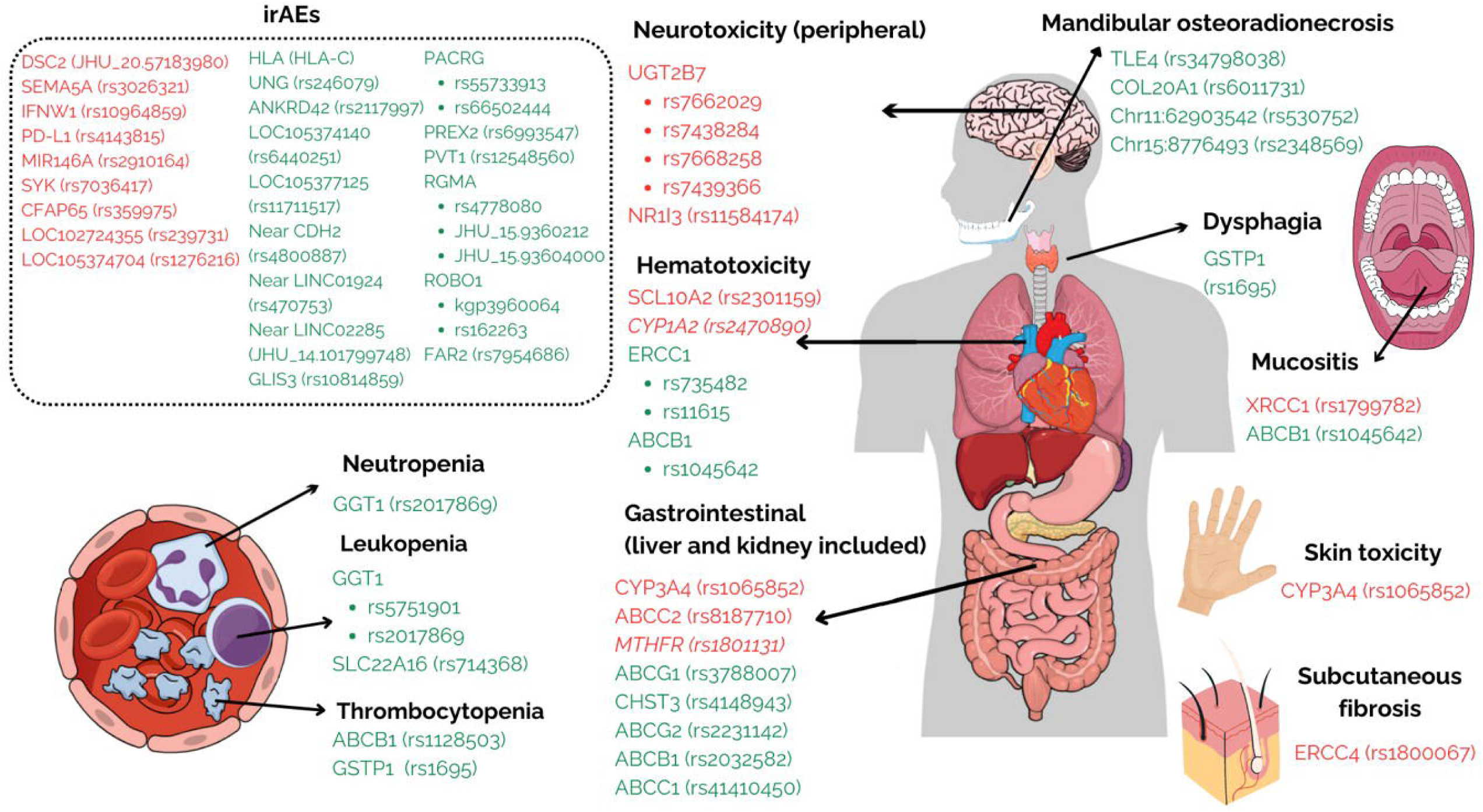

### SNVs associated with Immune-related Adverse Events (irAEs)

IrAEs usually developed mainly in non-small cell lung cancer (NSCLC) and melanoma within three months of ICI therapy and awere related to treatment with glucocorticoids, although some cases become chronic and require lifelong care (26,27). Among ICI treatments, CTLA-4 inhibitors had shown higher incidence of irAEs (∼70%) than PD-1/PD-L1 inhibitors (∼40%) (28). IrAEs, like skin toxicity, colitis, hepatitis, pneumonitis, and myocarditis occurred in 15–90% of the trials (30). Management of ICI-treated patients included corticosteroids or stopping therapy, with variations according to severity (31).

IrAEs mimic autoimmune diseases, such aslike ICI-induced pancreatitis, even though their autoimmune nature is not fully demonstrated (32). Although the underlying mechanisms are unclear, autoreactive T cells, autoantibodies, and cytokines may play important roles in the development of irAEs (29). In fact, patients with autoimmune diseases face serious challenges in ICI treatment, such as increased gastrointestinal toxicity in patients with inflammatory bowel disease (40% vs. 11%) (33). In addition, a meta-analysis of 613 fatal ICI events reported colitis (37-70%), pneumonitis (35%), and myocarditis (25%), with fatality rates of 0.36% for PD-1, 0.38% for PD-L1, and 1.08% for CTLA-4 (34).

Autoimmunity is classically linked to several HLA genes, and numerous studies have focused on HLA and irAEs. Luo et al. found that the *HLA* SNV rs9268543 was significantly associated with thyroid-related irAEs (p = 7.5×10−7) and linked to various autoimmune disorders in NSCLC (35), while *HLA-DR15, HLA-B52*, and *HLA-Cw12* variants (36) were more associated with pituitary irAEs. In contrast, some studies found no correlation between HLA genes and irAEs (Iafolla et al. (37) and Ali et al. (38)).

Other studies have focused on genomic regions different from *HLA* and some of them have determined several irAE-associated genetic variants, like *ANO5*, *IL1RL1*, *CFAP36, PNPT1* (39), *SYK* (40), *UNG*, *IFNW1*, *CTLA4*, *PD-L1*, *IFNL4* (41), *APP* (42), and *IL22RA1* (43).

Interestingly, Abdel-Wahab et al. (44) also found 12 SNVs, including *GABRP* rs11743438, rs11743735, and *DSC2* JHU_20.57183980, which were significantly associated with an increased risk of irAEs, and 18 variants related to a reduced risk of irAEs, such as *LOC105377125* rs11711517, JHU_15.93602126, and JHU_15.93604000 (44). Taylor et al. found *IL-7* rs16906115 to be a risk allele for irAEs after ICI treatment for melanoma, which was linked to an increased IL-7 (Interleukin 7) production by CD19+ B lymphocytes. However, even if this allele enhances the risk of irAEs due to *IL-7* association with T lymphocyte hyperexpansion, patients with this inflammatory profile exhibit better survivorship (45). In addition, Groha et al. identified the SNV rs75824728 within an intron in *IL22RA1* gene on chromosome 1p36 as a significant risk factor for all-grade irAEs in a study across several cancer types (43). Of note, SNVs linked to a higher risk of severe irAEs in ICI-treated patients were also found in genes coding for microRNAs, like rs2910164 in *MIR-146A* gene (46). Table 1 summarises the recently identified genetic studies and variants associated with novel toxicological irAEs.

### SNVs associated with toxicities in other oncology treatments

Genetic studies have primarily focused on identifying variations associated with adverse reactions to RT or chemotherapy. Toxicity was mainly reported on barrier organs, such as the skin, gastrointestinal tract, respiratory tract, and liver. Table 2 summarises the reviewed studies, including cancer type, toxicity, genes, location, and methodology. Key genes linked to toxicity include transporter genes, such as *ABC*, DNA repair genes, such as *XRCC*, and transcription factors, such as *NFE2L2*.

#### Dermatologic toxicity

Cancer treatments often cause dermatological toxicities, including rash, dryness, itching, nail changes, and hyperpigmentation. These skin conditions can significantly affect patients’ quality of life, self-image, and mental health, potentially resulting in treatment interruptions or dose reductions (47). Gupta et al. reported several genetic variants associated with the development of RT-related severe grade 3 subcutaneous fibrosis in spectrum HNSCC, including *XRCC3* rs861539 (p=0.015), *ERCC4* rs1800067 (p=0.012), *XRCC5* rs828907 (p=0.038), and *TGFβ1* rs1982073 (p=0.020) (48). Likewise, Raturi et al. found a significant association between *XRCC1* rs1799782 polymorphism and chemotherapy-related skin toxicity (p=0.01) in patients with advanced laryngeal squamous cell carcinoma (49). Conversely, Gudur et al. found no association between *XRCC1*, *XRCC2*, *XRCC3*, *XPC*, *XPD*, and *XPG* genes and RT-derived toxicity in HNSCC patients (50,51).

In addition, patients with nasopharyngeal cancer (NPC) treated with RT also reported skin toxicity associated with variants on chromosome 2q14 (rs6711678, rs4848597, rs4848598, and rs2091255) (52). In line with this, Cargnin et al. found an association between *TP53* rs1042522 and radiation-induced late skin toxicity in patients with breast cancer (BC) (p=0.028) (53), and Kwok et al. identified C*YP2D610* rs1065852 as a risk factor for gefitinib-induced skin toxicity in NSCLC (p = 0.050) (54).

#### Haematological toxicity

Haematopoietic toxicity involves a reduced production of rapidly dividing cells, like blood cell progenitors, which are essential for haematopoietic cell renewal and differentiation (55), and has been addressed mainly in NSCLC and BC.

Studies on NSCLC found some genes carrying associated variants with high risk of leukopenia induced by chemotherapy, like *SVIL* (p = 2.48E-06), *EFCAB2* (p = 4.63E-06) (56) and *ABCC1* rs41410450 (p = 0.04) (57). A higher risk of anaemia was also found in patients carrying *ABCC1* rs41410450 (p = 0.03) and *ABCC2* rs8187710 (p = 0.03) variants (57), as well as associations between thrombocytopenia and variants like *JMJD1C* rs34491125 (P = 9.07×10^-5) and *DOCK8* rs10491684 (P = 1.95×10^-4) (58). In addition, Xie et al. found an association between *XRCC1* rs25487 and a higher risk of severe lymphopenia in patients with NSCLC (p = 0.018) under chemoradiotherapy (59). Interestingly, Björn et al. developed a myelosuppressive toxicity prediction model for NSCLC patients treated with gemcitabine/carboplatin based on whole-genome sequencing data using a machine-learning LASSO model with 62 genetic variants (60). Conversely, De Troia et al. found *ABCB1* rs1045642 linked to a lower haematological toxicity risk in platinum-based chemotherapy (p = 0.01) (61).

Regarding BC, a significant number of studies have found genetic variants linked to protection against haematological toxicity. Ferracini et al. found that the *GSTP1* (rs1695) G allele reduced the risk of thrombocytopenia (p = 0.03) and anaemia (p = 0.01), whereas rs1128503 *ABCB1* was associated with an increased risk of thrombocytopenia (p = 0.03) (62). Similarly, Sun et al. reported that *GGT1* rs2017869 and rs5751901 were significantly associated with a lower risk of grade 2 or greater neutropenia (p = 0.036) and leukopenia (p = 0.024) (63). Likewise, Ebaid et al. found that *SLC22A16* rs714368 was significantly associated with a lower risk of neutropenia (p = 0.01, p =0.036) and leukopenia (p = 0.001, p = 0.024) (64). On the other hand, no significant association was found between *CBR1* rs20572 variant and DOX-induced haematological toxicity in BC patients (64).

Moreover, Duran et al. found an association between *ERCC1* rs11615 (p = 0.0066), *ERCC1* rs735482 (p = 0.0204), and *ERCC4* rs1799801(p = 0.0286) variants and a reduced grade 2-3 haematological toxicity in HNSCC (65). In addition, De Marchi et al. identified *SLC10A2* rs2301159 (OR = 25.52, p = 0.04) and *CYP1A2* rs2470890 (OR = 10.79, p = 0.038) as significant risk factors for haematological toxicity in Head and Neck squamous cell carcinoma (HNSCC) (66). In addition, Guo et al. found a variant in *Linc00312* rs15734 gene associated with severe leukopenia (p = 0.029) and rs12497104 with a reduced risk of thrombocytopenia (p = 0.030) in a study of young patients with NPC undergoing chemoradiotherapy (67). Finally, *ATG10* rs10514231 and *ATG16L2* rs10898880 were significantly associated with grade 3–4 myelosuppression in NPC patients undergoing RT (p < 0.001 and p = 0.010, respectively) (68).

Regarding colorectal cancer, *MTHFR* rs1801133 and rs1801131 were linked to chemotherapy-induced anemia (p = 0.005 and, p =0.002, respectively) and thrombocytopenia (p < 0.001 and p=0.03, respectively), as well as *DPYD* rs3918290 (p = 0.035 and p = 0.039, respectively) (69), whereas *MTHFR* rs1801133 was associated with neutropenia (p < 0.001) (70).

In 5-fluorouracil-treated oesophageal cancer, significant correlations were observed between *ABCC2* rs12762549 and *ABCB1* rs1045642 with severe neutropenia (71), and *ABCC2* rs717620 with grade 3–4 haematological toxicity (72). Moreover, methotrexate-induced neutropenia was linked to *ATF5* rs8667 and *SHMT1* rs1979277 variants in osteosarcoma patients (73). Lastly, Urtasun et al. identified multiple SNVs associated with haematological toxicities in infant cancer: *MTHFR* rs1801131 was linked to anemia; *C8orf34* rs1517114 and XPC rs2228001 were associated with neutropenia; and *ABCB1* rs1045642, *TNFRSF11B* rs2073618, *CYP2B6* rs4802101, and *SOD2* rs4880 were associated with thrombocytopenia (74).

#### Gastrointestinal toxicity

Gastrointestinal toxicity is a prevalent and often severe side effect of cancer treatment that manifesting as nausea, vomiting, diarrhoea, constipation, mucositis, and abdominal pain (75). Cancer treatment-induced mucositis has been predominantly studied in patients with HNSCC and nasopharyngeal carcinoma (NPC). Regarding NPC, variants in *XRCC1* rs1799782 (p = 0.01) (77), *ATG10* rs10514231 (p = 0.001), and *ATG16L2* rs10898880 (p = 0.001) (68) were associated with high oral mucositis in patients treated with RT, while *XRCC1* rs1799782 was associated to acute radiation-induced oral mucositis (p = 0.01) in NPC patients receiving chemoradiotherapy (49). Regarding RT and HNSCC, Gong et al. found an association between carrying the *XRCC1* rs25487 (p = 0.003) variant to an increased risk of severe mucositis (78). Moreover, a genome-wide association study (GWAS) yielded association of *TNKS* rs117157809 (p = 6.33 × 10⁻⁶), *VTA1* rs9484606 (p = 2.98 × 10⁻⁶), and *PER4* rs16876733 (p = 3.05 × 10⁻⁶) with an increased risk of oral mucositis (79), in addition to *STING1* rs28419191, rs10875554, and *DNAJC18* rs1131769 variants (80). Another meta-GWAS analysis performed by Naderi et al. found a top hit associated with mucositis near the *ZNF573* gene, rs73039018, which has transcription factor activity and might be linked to dysregulation of the inositol phosphate pathway mediating RT treatments (81). In addition, a link between mucositis and the rs67376798, rs55886062, and rs56038477 variants in *DPYD* was found in patients with advanced oropharyngeal carcinoma undergoing treatment with carboplatin / 5-FU (p = 0.0063) and RT (82). Regarding other tumours, Minnai et al. identified an association between *the ABCB1* rs1045642 variant and mucositis induced by doxorubicin and cisplatin in osteosarcoma (73).

Regarding other gastrointestinal toxicities *DPYD* (rs67376798, rs55886062, and rs56038477), which were linked to dysphagia (p = 0.019) in a study of oropharyngeal carcinoma treated with carboplatin and 5-FU (82). Additionally, a GWAS-derived variant, rs1061660 (located near *MTFP1*), was associated with dysphagia, while *TNFRSF21* rs6458543 was linked to xerostomia in patients with HNSCC (81). Conversely, *GSTP1* was significantly associated with a lower risk of severe dysphagia (65) in HNSCC patients treated with platinum-based chemoradiotherapy. Lastly, *ABCC2* rs8187710 (p = 0.001) and *MTHFR* rs1801131 (p = 0.015) were associated with an increased risk of gastrointestinal toxicity, whereas *ABCG1* rs3788007 and *CHST3* rs4148943 were associated with a decreased risk (p = 0.019 and p = 0.021, respectively) (66) in patients treated with cisplatin and paclitaxel. A high prevalence of *SYT8* rs584547 (P = 1.27 × 10^-6) (52) was observed in NPC patients treated with RT.

In addition, *CYP3A4*\*1/*1G (rs2242480) may increase gastrointestinal toxicity in patients with NSCLC treated with gefitinib (p = 0.006) (54). Likewise, Ishikawa et al. linked various polymorphisms, such as *EGFR* rs2293348 and rs4947492, with severe osimertinib-induced adverse events, notably anorexia, and *ABCG2* rs2231137 and *ABCB1* rs1128503 with grade ≥ 2 AEs (83). However, *ABCG2* rs2231142 was associated with a lower incidence of diarrhea (57). In a BC study, *AURKA variant,* rs8173 was linked to chemotherapy-induced gastrointestinal toxicity (p = 0.029) (84). Finally, Aoullay et al. found that grade III/IV nausea was more common in colorectal cancer patients carrying *ABCC2* rs1885301 (p = 0.004), and the absence of diarrhoea was associated with *ABCC2* rs2273697 in patients with pancreatic cancer (85).

#### Neurological toxicity

Cancer treatments frequently cause neurological toxicities that affect patients’ quality of life and treatment outcomes (86).

Several studies have identified a number of genetic variants associated with chemotherapy-induced peripheral neuropathy in patients with BC; for example, *EPHA5* rs7349683 (p = 0.008) and *EPHA8* rs209709 (p = 0.017) were associated with increased paclitaxel-induced neuropathy risk (87), which was replicated by Marcath et al., which confirmed the results of *EPHA5* rs7349683 (p = 0.007) (88). Likewise, Tolaney et al. linked *LOC154449* rs3012437 (p = 0.024) to a higher risk of paclitaxel-induced neuropathy in HER2-positive patients receiving adjuvant paclitaxel and trastuzumab (89). However, other studies have identified various genotypes that offer protection against neurotoxicity in BC patients undergoing neoadjuvant chemotherapy like *GGT1* rs8135987 (p = 0.042) (63). Regarding taxane-based treatment of BC, a large study of 21 SNVs across 13 genes revealed significant correlations between grade 2-3 neuropathy and *UGT2B7* variants rs7438284, rs7662029, rs7439366, rs7668258, and *NR1I3* rs11584174 (90). Moreover, Chua et al. found an association between *S1PR1* rs74497159, *C9orf106* rs77526807, *SLITRK1* rs17076837, *KLHL1* rs61963755, *FGD4* rs10771973, *ZFPM2* rs3110366, rs2342780, and rs2342791 SNVs through GWAS analyses of BC patients treated with microtubule-targeting agents (91). In addition, *SCN9A* rs13017637 was found to be a significant predictor of grade 2 or higher taxane-induced neuropathy (p = 0.0050) in Japanese patients with breast and ovarian cancer (92). Additionally, *ABCB1* rs1045642 was associated with a higher risk of developing grade 2 and 3 neurotoxicity (p = 0.03) in chemotherapy-treated patients with ovarian cancer (62).

Moreover, the GG genotype of *TRPV1* rs879207 (p = 0.012) was associated with a 5-fold increased risk of severe neuropathy NSCLC in patients receiving platinum-based treatment (93), *MTHFR* rs1801133 was associated with neuropathy in patients with metastatic colorectal cancer under chemotherapy (p = 0.02) (70), and *ABCG2* rs2231137 was associated with an increased risk of neutropenia related to palbociclib treatment (94).

Lastly, *GPR68* rs17020773 was associated with peripheral neuropathy (p = 2.03 × 10^-8) in patients with taxane-treated triple-negative BC, renal cell carcinoma (RCC), NSCLC, small cell lung cancer, bladder cancer, ovarian cancer, and melanoma (95).

#### Other toxicities

We found numerous studies linking other genes with toxicities that did not fit within previous categories.

In patients with HNSCC, Fujikawa et al. linked the *NFE2L2* rs6721961 variant to early cisplatin-derived ototoxicity (p = 0.03) (96) and found significant associations between the rs34402524 *TET2*, rs149936944 *GEN1*, and rs35831183 *NCOR2* variants with long-term RT-derived toxicity (97). *TLE4* rs34798038, *COL20A1* rs6011731, and Chr11:62903542 rs530752 genetic variants were linked to the absence of mandibular osteoradionecrosis in HNSCC under RT (98), and *DPYD* rs67376798, rs55886062, and rs56038477 were associated with aspiration pneumonia (p = 0.00065) in oropharyngeal carcinoma treated with carboplatin and 5-FU(82).

Concerning NSCLC, severe radiation-induced toxicity was associated to *XRCC1* rs25487, *XRCC2* rs3218556, and *XPD* rs13181 (p < 0.05) (77), while platinum-based toxicity was lin ked to *ERCC5* SNV rs4771436, rs11069498, rs4150330 (99) and *VEGF* rs1570360, which were specifically associated with higher grade 3-4 toxicities (p = 0.004) (100). Moreover, *ABCB1* rs2032582 and *ABCC1* rs41410450 were associated with reduced nephrotoxicity, whereas rs41410450 *ABCC1*, rs1128503 *ABCB1,* and rs2032582 *ABCB1* were associated with hepatotoxicity (57) in patients undergoing platinum-based doublet chemotherapy. Similarly, CYP2D6*41 (rs16947) may increase hepatotoxicity in gefitinib patients with NSCLC (p = 0.04) (54). Nonetheless, no association between *NRF2* rs6721961 and toxicity was found in gemcitabine-treated patients with lung cancer (101).

Regarding BC, patients undergoing RT, *ANOS1* rs188287402, *LOC124901047* rs12657177, and *PAX7* rs643644 were found to be associated with nipple retraction (p = 2.80 × 10^-8), breast edema (p ≤ 1.23 × 10^-9), and arm lymphedema (p = 3.54 × 10^-8), respectively (102). *eNOS* rs2070744 was associated with proteinuria with bevacizumab (103), while *OPG* rs2073618 was associated with musculoskeletal symptoms following aromatase inhibitor treatment (p = 0.004) (104). Sun et al. linked *GGT1* rs8135987 with increased gamma-glutamyl transferase as a sign of hepatotoxicity in BC patients undergoing neoadjuvant chemotherapy (63). In contrast, the co-occurrence of *CYP19A1* rs10046 and rs749292 was linked to a reduced 10-year cumulative skeletal event in postmenopausal BC patients (p = 0.033) (105), while Hertz et al. found no link between *TCL1A* rs11849538 and musculoskeletal symptoms caused by aromatase inhibitors (104). Three studies were reviewed in relation to BC and cardiac toxicity: Velasco-Ruiz et al. found that *POLRMT* rs62134260 was linked to cardiotoxicity in BC patients receiving anthracycline-based chemotherapy (p = 7.10 × 10−6) (106), and Liu et al. associated cardiac toxicity with *SLC28A3* rs885004 (p = 0.010) (107). In addition, Conte et al. associated rs727479 with cardiovascular events in patients receiving aromatase inhibitors (p = 0.026) (105).

Regarding metastatic colorectal cancer, *MTHFR* rs1801133 was found to be associated with diarrhea (p = 0.005) and hand-foot syndrome (p = 0.013) (70), whereas homozygous carriers of *SULT2A1* rs182420 variant had a higher risk of toxicity due to palbociclib (p = 0.042) (94).

In addition, Naderi et al. identified SNVs linked to acute standardised total average toxicity (STAT_acute_) with *DPPA4* rs142667902 across prostate, HNSCC, BC, and lung cancer (108). Finally, regarding pediatric patients, Meijer et al. linked *TCERG1L* rs893507 to hearing loss in cisplatin-treated cancer patients (p = 5.3 × 10^-10) (109).

## Discussion

This review examines the last five years (December 2019–July 2024) of research on genetic variants associated with treatment-related toxicities in solid tumours. The most frequently studied cancer types were breast cancer (18 studies), non-small cell lung cancer (15 studies), and head and neck cancers (13 studies). Regarding treatment modalities, 39 studies focused on chemotherapy-, 12 on immunotherapy-, 14 on radiotherapy-, and 5 on chemoradiotherapy-related toxicity. A total of 19 genes were implicated in a broad spectrum of toxicities, such as peripheral neuropathy, as well as haematological and gastrointestinal adverse effects. Conversely, 111 genes were associated with one type of toxicity (Tables S1 and S2). The analisys of odds ratios revealed that 68 genetic varints were linked to an increased risk (OR>1), while only 25 had a protective role (OR<1). Interestingly, SNVs in nine genes (*ABCB1, ABCC1, ABCG2, ERCC4, ERCC5, GGT, HLA, Linc00312,* and *XRCC1*) were associated with both risk and protection.

Alleles in genes such as *GSTP1, ABCG1, CHST3, ABCB1, ERCC1, GGT1, SLC22A16, HLA, UNG, COL20A1, and TLE4,* which are predominantly involved in cellular transport, detoxification, DNA repair, extracellular matrix structure, or immune tolerance, showed a protective effect in the development of treatment associated toxicities. Conversely, alleles in genes like *CYP3A4, ABCC2, MTHFR, CYP12, SLC10A2, DSC2, IFNW1, MIR146A, PD-L1, SEMA5A, SYK, XRCC1, NFE2L2, NR1I3, UGT27B7* and *ERCC4,* were associated with the development of toxicities, displayed a broad functional diversity, and played key roles in drug and xenobiotic metabolism, gene regulation, cellular signalling, and immune response.

In view of the technological advancements in the field, future research should move beyond single-gene associations or GWAS to investigate the combined effects of multiple exonic genetic variants, as well as their interactions with other molecular pathways influencing treatment response. Expanding the use of next-generation sequencing (NGS) could accelerate the identification of toxicity-related markers and enhance the potential of pharmacogenomics to personalise cancer treatments. This is particularly relevant in light of the increasing use of combination therapies, such as immunotherapy alongside traditional treatments, which may lead to enhanced toxicity risks (110–112). Developing NGS-based panels that incorporate high-risk and highly protective genetic variants, as categorised in this review (OR > 6, OR < 0.3), across multiple cancer types, and the development of machine-learning models, as biorg et al. (60) suggested, could significantly enhance treatment planning and risk stratification. The integration of pharmacogenomic testing into routine clinical practice could enhance patient stratification, minimise toxicity, and optimise the therapeutic efficacy. Furthermore, integrating pharmacogenomic data with other ‘’ omics technologies, such as transcriptomics, proteomics, and metabolomics, could provide a more comprehensive approach to understanding and predicting treatment-related toxicities. However, the use of diverse methodologies, such as NGS, MassArray, and PCR-RFLP, together with high variability between experimental designs, toxicity assessment, and lack of replication studies complicates standardisation, which is key to translating these findings into clinical practice. Moreover, given the known genetic variability in drug metabolism and immune responses, large-scale, standardised multicentre studies to validate pharmacogenomic biomarkers would ensure that pharmacogenomic advancements benefit all patient populations and are clinically reliable.

All in all, common efforts should point to standardization to bring the field closer to fully personalised oncology, where genetic insights drive clinical decision-making, improving patient outcomes, and minimizing adverse effects.

## Conclusion

Pharmacogenomics presents a powerful opportunity for personalised cancer treatment, allowing the identification of genetic variants linked to therapy-induced toxicities and enabling better patient stratification. However, to translate these findings into routine clinical practice, future research must prioritise standardised methodologies and large-scale validation studies to ensure broad applicability and clinical reliability.

## Supporting information

Table S1. Novel toxicological SNVs included

igure S1. Protective (OR<0.3) (A) and B) risky (OR>6) (B) SNVs forest plots

## Data Availability

All data produced in the present work are contained in the manuscript

## ABBREVIATIONS

5-FU: 5-Fluorouracil
ABC: ATP-Binding Cassette
ABCB1: ATP-Binding Cassette Subfamily B Member 1
ABCC1: ATP-Binding Cassette Subfamily C Member 1
ABCC2: ATP-Binding Cassette Subfamily C Member 2
ABCG1: ATP-Binding Cassette Subfamily G Member 1
ABCG2: ATP-Binding Cassette Subfamily G Member 2
ANO5: Anoctamin 5
ANOS1: Anosmin 1
APP: Amyloid Precursor Protein
ATF5: Activating Transcription Factor 5
ATG10: Autophagy Related 10
ATG16L2: Autophagy Related 16 Like 2
AURKA: Aurora Kinase A
BC: Breast cancer
BRAF: v-raf murine sarcoma viral oncogene homolog B1
C8orff34: Chromosome 8 Open Reading Frame 34
CBR1: Carbonyl Reductase 1
CD19+: Cluster of Differentiation 19
CFAP36: Cilia and Flagella Associated Protein 36
CHST3: Carbohydrate Sulfotransferase 3
COL20A1: Collagen Type XX, Alpha 1
CRT: Chemoradiotherapy
CTLA-4: Cytotoxic T-Lymphocyte-Associated protein 4
CYP19A1: Cytochrome P450 Family 19 Subfamily A Member 1
CYP1A2: Cytochrome P450 Family 1 Subfamily A Member 2
CYP2D6: Cytochrome P450 Family 2 Subfamily D Member 6
CYP2D610: Cytochrome P450 Family 2Subfamily D Member 6
CYP3A4: Cytochrome P450 Family 3 Subfamily A Member 4
DALY: Disability-adjusted life years
DNAJC18: DnaJ Heat Shock Protein Family (Hsp40) Member C18
DOCK8: Dedicator of Cytokinesis 8
DOX: Docetaxel
DPPA4: Developmental Pluripotency Associated 4
DPYD: Dihydropyrimidine Dehydrogenase
DSC2: Desmocollin 2
EFCAB2: EF-Hand Calcium Binding Domain 2
EGFR: Epidermal Growth Factor Receptor
eNOS: Endothelial Nitric Oxide Synthase
EPHA5: Ephrin Type-A Receptor 5
EPHA8: Ephrin Type-A Receptor 8
ERCC4: Excision Repair Cross-Complementation Group 4
ERCC5: Excision Repair Cross-Complementation Group 5
FGD4: FYVE, RhoGEF and PH Domain Containing 4
GABRP: Gamma-Aminobutyric Acid Type A Receptor Pi Subunit
GEN1: Gene 1
GGT1: Gamma-Glutamyltransferase 1
GPR68: G Protein-Coupled Receptor 68
GSTP1: Glutathione S-Transferase Pi 1
GWAS: Genome-Wide Association Study
HER2: Human Epidermal growth factor Receptor 2
HLA: Human Leukocyte Antigen
HNSCC: Head and neck squamous cell cancer
ICI: Immune checkpoint inhibitors
IFNL4: Interferon Lambda 4
IFNW1: Interferon Omega 1
IL1RL1: Interleukin 1 Receptor-Like 1
IL22RA1: Interleukin 22 Receptor Alpha 1
IL-7: Interleukin 7
irAEs: Immune-related adverse events
JMJD1C: Jumonji Domain Containing 1C
KLHL1: Kelch Like Family Member 1
Linc00312: Long Intergenic Non-Protein Coding RNA 312
MIR-146A: MicroRNA 146A
MTFP1: Mitochondrial Fission 1 Protein
MTHFR: Methylenetetrahydrofolate Reductase
NCOR2: Nuclear Receptor Corepressor 2
NFE2L2/NRF2: Nuclear Factor Erythroid 2-Like 2
NFE2L2: Nuclear Factor, Erythroid 2-Like 2
NGS: Next Generation Sequencing
NPC: Nasopharyngeal cancer
NR1I3: Nuclear Receptor Subfamily 1 Group I Member 3
NSCLC: Non-small-cell lung cancer
OPG: Osteoprotegerin OR odds ratio
PAX7: Paired Box Gene 7
PD-1: Programmed Cell Death Protein 1
PD-L1: Programmed Death-Ligand 1
PER4: Period Circadian Regulator 4
PNPT1: Polyribonucleotide Nucleotidyltransferase 1
POLMRT: Poly (ADP-Ribose) Polymerase Family Member
PPM: Personalised presicion medicine
RT: Radiotherapy
RCC: Renal Cell Carcinoma
S1PR1: Sphingosine-1-Phosphate Receptor 1
SCN9A: Sodium Channel Voltage-Gated Type IX Alpha Subunit
SHMT1: Serine Hydroxymethyltransferase 1
SLC10A2: Solute Carrier Family 10 Member 2
SLC22A16: Solute Carrier Family 22 Member 16
SLC28A3: Solute Carrier Family 28 Member 3
SLITRK1: SLIT and NTRK-like Family Member 1
SNV: Single nucleotide variants
SOD2: Superoxide Dismutase 2
STING1: Stimulator of Interferon Genes 1
SULT2A1: Sulfotransferase Family 2A Member 1
SVIL: Supervillin
SYK: Spleen Tyrosine Kinase
TCERG1L: Transcription Elongation Regulator 1-Like
TET2: Ten-Eleven Translocation 2
TGFβ1: Transforming Growth Factor Beta 1
TLE4: Transducin-Like Enhancer of Split 4
TNFRSF11B: Tumor Necrosis Factor Receptor Superfamily Member 11B
TNFRSM21: Tumor Necrosis Factor Receptor Superfamily Member 21
TNKS: Tankyrase
TP53: Tumor Protein p53
TRPV1: Transient Receptor Potential Cation Channel Subfamily V Member 1
UGT2B7: UDP Glucuronosyltransferase Family 2 Member B7
UNG: Uracil-DNA Glycosylase
VEGF: Vascular Endothelial Growth Factor
VTA1: Vesicle Trafficking 1
XPC: Xeroderma Pigmentosum, Complementation Group
C XPD: Xeroderma Pigmentosum, Complementation Group
D XPG: Xeroderma Pigmentosum, Complementation Group
G XRCC: X-Ray Repair Cross-Complementing
XRCC1: X-Ray Repair Cross-Complementing 1
XRCC2: X-Ray Repair Cross-Complementing 2
XRCC3: X-Ray Repair Cross-Complementing 3
XRCC5: X-Ray Repair Cross-Complementing 5
ZFPM2: Zinc Finger Protein, FOG Family Member 2
ZNF573: Zinc Finger Protein 573

## Acknowledgements

Not aplicable

## Funding

Proy_Excel_01002

### Author contributions

Isabel Barragán, Javier Oliver, Elisabeth Pérez-Ruiz, Alejandro Escamilla, and Antonio Rueda-Domínguez conceptualization, methodology, data curation, writing–original draft, writing–review and editing, supervision, project administration Andrea Gonzalez-Hernandez Data Curation, Writing - Original Draft, Cecilia A. Frecha, Felipe Vaca-Paniagua, Sandra Perdomo, Antonio Rueda-Domínguez, Writing - Review & Editing, Supervision

### Ethics declarations

Ethics approval and consent to participate Not applicable.

### Consent for publication

All authors provided consent for publication.

