## Supplementary material for "From Genes to Personalized Cancer Care: A Systematic Review of Toxicity-Associated Genetic Variants in Solid Tumor Treatments": Table S1. Novel toxicological SNVs included

### Appendix A. Supplementary material

**Table S1**. Novel toxicological SNVs included

| Toxicity | Study | Genes | Variants | OR | pvalue |
|---|---|---|---|---|---|
| irAEs | Abdel-Wahab et al. 2021 (44) | *AGPS* | JHU_2.179128562 | 4.6 | 8.29 × 10-5 |
| | | *ANKRD42* | rs2117997 | 0.22 | 2.44 × 10-5 |
| | | *BAZ2B* | rs56328422 | 4.2 | 4.14 × 10-5 |
| | | *CFAP65* | rs359975 | 8.5 | 9.41 × 10-5 |
| | | *DSC2* | JHU_20.57183980 | 6.9 | 8.85 × 10-6 |
| | | *FAR2* | rs7954686 | 0.16 | 3.98 × 10-5 |
| | | *GABRP* | rs11743438<br>rs11743735 | 4.3<br>4.5 | 5.56 × 10-6<br>8.34 × 10-6 |
| | | *GLIS3* | rs10814859 | 0.24 | 5.88 × 10-5 |
| | | *LOC102724355* | rs239731 | 11.3 | 8.56 × 10-5 |
| | | *LOC105374140* | rs6440251 | 0.13 | 5.15 × 10-5 |
| | | *LOC105374704* | rs1276216 | 6.4 | 8.18 × 10-5 |
| | | *LOC105377125* | rs11711517 | 0.16 | 2.39 × 10-6 |
| | | *Near CDH2* | rs4800887 | 0.25 | 5.74 × 10-5 |
| | | *Near DCBLD2* | rs2062059 | 0.28 | 7.37 × 10-5 |
| | | *Near LINC01924* | rs470753 | 0.16 | 1.65 × 10-5 |
| | | *Near LINC02285* | JHU_14.101799748 | 0.11 | 5.20 × 10-5 |
| | | *OSBPL6* | rs919682 | 4.6 | 8.29 × 10-5 |
| | | *PACRG* | rs55733913<br>rs66502444 | 0.16<br>0.16 | 3.98 × 10-5<br>9.11 × 10-5 |
| | | *PREX2* | rs6993547 | 0.16 | 9.11 × 10-5 |
| | | *PVT1* | rs12548560 | 0.28 | 8.95 × 10-5 |
| | | *RGMA* | rs4778080<br>JHU_15.9360212<br>JHU_15.93604000 | 0.24<br>0.24<br>0.24 | 1.92 × 10-5<br>8.89 × 10-6<br>8.89 × 10-6 |
| | | *ROBO1* | kgp3960064<br>rs162263 | 0.28<br>0.24 | 6.76 × 10-5<br>5.88 × 10-5 |
| | | *SEMA5A* | rs3026321<br>JHU_5.9269447 | 19.8<br>3.5 | 6.31 × 10-5<br>8.61 × 10-5 |
| | | *Near ZPLD1* | rs35807769 | 4.9 | 6.07 × 10-5 |
| | Ali et al. 2019 (38) | *HLA* | HLA-DRB1*11:01 | 4.53<br>3.94 | 2.1 × 10-3<br>1.7 × 10-2 |

| | | | | | |
|---|---|---|---|---|---|
| | | | HLA-DQB1*03:01 | | |
| | Ferguson et al. 2023 (40) | *SYK* | rs7036417 | 7.46 | 1.43 × 10-04 |
| | Groha et al. 2022 (43) | *IL22RA1* | rs75824728 | HR 1.8 | 3.5 × 10-8 |
| | | *Near LINC02484* | rs113861051 | 2.0 | 1.2 × 10-8 |
| | Iafolla et al. 2021 (37) | *HLA* | HLA-A<br>HLA-B<br>HLA-C | HLA-C 0.28 | 4 × 10-2 |
| | Luo et al. 2021 (35) | *HLA* | rs9268543 | NA | 7.5 × 10-7 |
| | Marschner et al. 2020 (46) | *miR-146A* | rs2910164 | 6.78 | 4 × 10-3 |
| | Montaudié et al. 2021 (39) | *ANO5* | rs7481951 | NA | 3.1 × 10-5 |
| | | *IL1RL1* | rs4988956 | NA | 9.2 × 10-2 |
| | | *PNPT1* | rs782637<br>rs782594<br>rs7882572 | NA | 4.1 × 10-8<br>3.4 × 10-7<br>3.8 × 10-7 |
| | | *CFAP36* | rs3762513 | NA | 6.6 × 10-6 |
| | Refae et al. 2019 (41) | *CTLA4* | rs3087243<br>rs11571302<br>rs7565213 | 3.19<br>4.06<br>3.73 | 4.8 × 10-2<br>1.8 × 10-2<br>2.6 × 10-2 |
| | | *IFNL4* | rs12979860 | 3.72 | 3.6 × 10-2 |
| | | *IFNW1* | rs10964859 | 6.08 | 1.4 × 10-2 |
| | | *PD-L1* | rs4143815 | 8.11 | 3 × 10-3 |
| | | *UNG* | rs246079 | 0.09 | 1 × 10-3 |
| | Taylor et al. 2022 (45) | *IL7* | rs16906115 | 2.24 | 4.6 × 10-2 |
| | Udagawa et al. 2022 (42) | *Near APP* | rs469490 | 5.15 | 2.97 × 10-7 |
| | Yano et al. 2020 (36) | *HLA* | HLA-DR15<br>HLA-Cw12<br>HLA-B52 | NA | 1.4 × 10-3<br>1.3 × 10-3<br>2.6 × 10-3 |
| Dermatologic | Cargnin et al. 2021 (53) | *TP53* | rs1042522 | 4.7 | 7 × 10-3 |
| | Gupta et al. 2021 (48) | *ERCC4* | rs1800067 | 23.14 | 1.2 × 10-2 |
| | | *TGFβ1* | rs1982073 | 4.61 | 2 × 10-2 |
| | | *XRCC3* | rs861539 | 2.23 | 1.5 × 10-2 |
| | | *XRCC5* | rs828907 | 3.06 | 3.8 × 10-2 |
| | Kwok et al. 2022 (54) | *CYP2D6* | rs1065852 | 3.37 | 5 × 10-2 |
| | Raturi et al. 2020 (49) | *XRCC1* | rs1799782 | NA | 1 × 10-2 |
| | Wang et al. 2022 (52) | *Chr 2: 2q14* | rs6711678<br>rs4848597<br>rs4848598<br>rs2091255 | 1.23<br>1.34<br>1.36<br>1.4 | 7.17 × 10-7<br>4.82 × 10-7<br>4.91 × 10-7<br>9.83 × 10-7 |

| | | | | | |
|---|---|---|---|---|---|
| Dysphagia | Duran et al. 2019 (65) | *GSTP1* | rs1695 | 0.2 | 5 × 10-4 |
| | Naderi et al. 2022 (81) | *MTFP1* | rs1061660 | NA | 9.9 × 10-6 |
| | Wang et al. 2022 (52) | *SYT8* | rs584547 | 4.2 | 2 × 10-3 |
| Gastrointestinal | Aoullay et al. 2023 (85) | *ABCC2* | rs1885301 | NA | 4 × 10-3 |
| | De Marchi et al. 2019 (66) | *ABCC2* | rs8187710 | 33.21 | 1 × 10-3 |
| | | *ABCG1* | rs3788007 | 0.04 | 1.9 × 10-2 |
| | | *CHST3* | rs4148943 | 0.05 | 2.1 × 10-2 |
| | | *MTHFR* | rs1801131 | 17.69 | 1.5 × 10-2 |
| | Desilets et al. 2022 (82) | *DPYD* | rs67376798<br>rs55886062<br>rs56038477 | RR 2.36 | 6.3 × 10-3 |
| | Kwok et al. 2022 (54) | *CYP3A4* | rs2242480 | 20 | 6 × 10-3 |
| | Macier-Cruz et al. 2023 (84) | *AURKA* | rs8173 | 1.6 | 2.9 × 10-2 |
| | Minnai et al. 2023 (73) | *ABCB1* | rs1045642 | 0.055 | 3.7 × 10-2 |
| | Naderi et al. 2022 (81) | *TNFRSF21* | rs6458543 | NA | 9.65 × 10-7 |
| | | *ZNF573* | rs73039018 | NA | 8.5 × 10-7 |
| | Raturi et al. 2020 (49) | *XRCC1* | rs1799782 | NA | 8 × 10-3 |
| | Schack et al. 2022 (80) | *DNAJC18* | rs1131769 | 2.2 | 7.6 × 10-8 |
| | | *STING1* | rs10875554 | 2.2 | 5.1 × 10-8 |
| | | | rs28419191 | 2.2 | 4.4 × 10-8 |
| | Sharma et al. 2022 (57) | *ABCG2* | rs2231142 | 0.25 | 4 × 10-2 |
| | Yang and Liu 2019 (68) | *ATG10* | rs10514231 | 1.95 | 1 × 10-3 |
| | | *ATG16L2* | rs10898880 | 1.56 | 1 × 10-3 |
| | Yang et al. 2020 (79) | *PER4* | rs16876733 | 1.95 | 3.05 × 10-6 |
| | | *TNKS* | rs117157809 | 3.72 | 6.33 × 10-6 |
| | | *VTA1* | rs9484606 | 1.7 | 2.98 × 10-6 |
| Hematological | Deligonul et al. 2021 (69) | *DYPD* | rs3918290 | NA | 3.5 × 10-2 |
| | De Marchi et al. 2019 (66) | *SLC10A2* | rs2301159 | 25.52 | 4 × 10-3 |
| | | *CYP1A2* | rs2470890 | 10.79 | 3.8 × 10-2 |
| | De Troia et al. 2019 (61) | *ABCB1* | rs1045642 | 0.2 | 1 × 10-2 |
| | Duran et al. 2019 (65) | *ERCC1* | rs11615<br>rs735482 | 0.29<br>0.28 | 6.6 × 10-3<br>2.04 × 10-2 |

| | | | | | |
|---|---|---|---|---|---|
| | | *ERCC4* | rs1799801 | 0.39 | 2.86 × 10-2 |
| | Ferracini et al. 2021 (62) | *GSTP1* | rs1695 | 0.17 | 1 × 10-2 |
| | Fujita et al. 2022 (72) | *ABCC2* | rs717620 | 1.77 | 3.8 × 10-2 |
| | Ramos-Esquivel et al. 2020 (70) | *MTHFR* | rs1801133<br>rs1801131 | 1.69<br>2.75 | 5 × 10-3<br>2 × 10-2 |
| | Sharma et al. 2022 (57) | *ABCC2* | rs8187710 | 4.24 | 3 × 10-2 |
| | | *ABCC1* | rs41410450 | 1.37 | 2 × 10-2 |
| | Urtasun et al. 2023 (74) | *MTHFR* | rs1801131 | 1.73 | NA |
| | Yang and Liu 2019 (68) | *ATG10* | rs10514231 | 2.08 | 1 × 10-3 |
| | | *ATG16L2* | rs10898880 | 1.56 | 1 × 10-3 |
| Leukopenia | Ebaid et al. 2024 (64) | *SLC22A16* | rs714368 | 0.18 | 1 × 10-3 |
| | Guo et al. 2022 (67) | *Linc00312* | rs15734 | 3.145 | 2.9 × 10-2 |
| | Sharma et al. 2022 (57) | *ABCC1* | rs41410450 | 1.88 | 4 × 10-2 |
| | Sun et al. 2023 (63) | *GGT1* | rs2017869<br>rs5751901 | 0.24<br>0.27 | 1.7 × 10-2<br>2.4 × 10-2 |
| | Svedberg et al. 2020 (56) | *SVIL* | rs3740003<br>rs10160013<br>rs1547169 | NA | 1.12 × 10-4<br>1.16 × 10-4<br>1.63 × 10-4 |
| | | *EFCAB2* | rs10927386<br>rs10927387 | NA | 2.52 × 10-6<br>2.52 × 10-6 |
| | Xie et al. 2020 (59) | *XRCC1* | rs25487 | HR 1.67 | 1.8 × 10-2 |
| Neutropenia | Ebaid et al. 2024 (64) | *SLC22A16* | rs714368 | 0.31 | 1 × 10-2 |
| | Minnai et al. 2023 (73) | *ATF5* | rs8667 | 0.11 | 3.4 × 10-2 |
| | | *SHMT1* | rs1979277 | 5.3 | 4 × 10-2 |
| | Nomura et al. 2020 (71) | *ABCB1* | rs1045642 | 2.1 | 2.7 × 10-2 |
| | | *ABCC2* | rs12762549 | 2.37 | 1.4 × 10-2 |
| | Ramos-Esquivel et al. 2020 (70) | *MTHFR* | rs1801133 | 2.27 | 1 × 10-3 |
| | Sun et al. 2023 (63) | *GGT1* | rs2017869<br>rs5751901 | 0.39<br>0.29 | 2.5 × 10-2<br>3.6 × 10-2 |
| | Urtasun et al. 2023 (74) | *C8orf34* | rs1517114 | 1.5 | NA |
| | | *XPC* | rs2228001 | 4.63 | NA |
| | Wang et al. 2024 (94) | *ABCG2* | rs2231137 | 4.14 | 5.2 × 10-2 |
| Thrombocytopenia | Björn et al. 2019 (58) | *DOCK8* | rs10491684 | NA | 1.95 × 10-4 |
| | | *JMJD1C* | rs34491125 | NA | 9.07 × 10-5 |

| | | | | | |
|---|---|---|---|---|---|
| | Deligonul et al. 2021 (69) | *DPYD* | rs3918290 | NA | 3.9 × 10-2 |
| | Ferracini et al. 2021 (62) | *ABCB1* | rs1128503 | 0.15 | 3 × 10-2 |
| | | *GSTP1* | rs1695 | 0.27 | 1 × 10-2 |
| | Guo et al. 2022 (67) | *Linc00312* | rs12497104 | 0.325 | 3 × 10-2 |
| | Ramos-Esquivel et al. 2020 (70) | *MTHFR* | rs1801133 | 1.91 | 1 × 10-3 |
| | Urtasun et al. 2023 (74) | *ABCB1* | rs1045642 | 1.7 | NA |
| | | *CYP2B6* | rs4802101 | 1.7 | NA |
| | | *SOD2* | rs4880 | 1.73 | NA |
| | | *TNFRSF11B* | rs2073618 | 1.77 | NA |
| Hepatotoxicity | Kwok et al. 2022 (54) | *CYP2D6* | rs16947 | NA | 4 × 10-2 |
| | Sharma et al. 2022 (57) | *ABCB1* | rs2032582<br>rs1128503 | 0.61<br>1.85 | 3 × 10-2<br>1 × 10-2 |
| | | *ABCC1* | rs41410450 | 2.06 | 2 × 10-2 |
| | Sun et al. 2023 (63) | *GGT1* | rs8135987 | 3.11 | 3.6 × 10-2 |
| Neuropathy | Arbitrio et al. 2019 (90) | *NR1I3* | rs11584174 | 6.133 | 1 × 10-3 |
| | | *UGT2B7* | rs7438284<br>rs7662029<br>rs7439366<br>rs7668258 | 6.34<br>6.34<br>6.34<br>6.34 | 2 × 10-3<br>2 × 10-3<br>2 × 10-3<br>2 × 10-3 |
| | Chua et al. 2020 (91) | *C9orf106* | rs77526807 | NA | 1.66 × 10-6 |
| | | *FGD4* | rs10771973 | NA | 2.15 × 10-6 |
| | | *KLHL1* | rs61963755 | NA | 1.88 × 10-6 |
| | | *S1PR1* | rs74497159 | NA | 3.62 × 10-7 |
| | | *SLITRK1* | rs17076837 | NA | 1.85 × 10-6 |
| | | *ZFPM2* | rs3110366<br>rs2342780<br>rs2342791 | NA<br>NA<br>NA | 1.07 × 10-6<br>2.06 × 10-6<br>2.53 × 10-6 |
| | Ciruelos et al. 2019 (87) | *EPHA5* | rs7349683 | HR 3.08 | 8 × 10-3 |
| | | *EPHA8* | rs209709 | HR 2.43 | 1.7 × 10-2 |
| | Khan et al. 2023 (95) | *GPR68* | rs17020773 | HR 1.82 | 4.39 × 10-6 |
| | Marcath et al. 2020 (88) | *EPHA5* | rs7349683 | NA | 7 × 10-3 |
| | Ramos-Esquivel et al. 2020 (70) | *MTHFR* | rs1801133 | 1.77 | 2 × 10-2 |
| | Sun et al. 2023 (63) | *GGT1* | rs8135987 | 0.39 | 4.2 × 10-2 |
| | Tolaney et al. 2019 (89) | *LOC154449* | rs3012437 | 2.1 | 2.4 × 10-2 |
| Other toxicities | Brooker et al. 2021 (98) | *COL20A1* | rs6011731 | 0.24 | 1.8 × 10-2 |

| | | | | | |
|---|---|---|---|---|---|
| | | *TLE4* | rs34798038 | 0.076 | 6 × 10-3 |
| | | *Chr11:62903542* | rs530752 | 0.24 | 4.6 × 10-2 |
| | Conte et al. 2023 (105) | *CYP19A1* | rs10046<br>rs749292 | NA<br>NA | 1.5 × 10-2<br>2.1 × 10-2 |
| | | *CYP19A1* | rs727479 | HR 0.23 | 4.8 × 10-2 |
| | Crucitta et al. 2019 (103) | *eNOS* | rs2070744 | NA | 4 × 10-3 |
| | Deichaite et al. 2022 (97) | *TET2* | rs34402524 | NA | 8 × 10-3 |
| | | *GEN1* | rs149936944 | NA | 2.7 × 10-3 |
| | | *NCOR2* | rs35831183 | NA | 8 × 10-3 |
| | Fujikawa et al. 2024 (96) | *NFE2L2* | rs6721961 | NA | 3 × 10-2 |
| | Hertz et al. 2021 (104) | *OPG* | rs2073618 | 3.33 | 4 × 10-3 |
| | Ishikawa et al. 2023 (83) | *ABCB1* | rs1128503 | NA | 3.8 × 10-2 |
| | | *ABCG2* | rs2231137 | NA | 8 × 10-3 |
| | | *EGFR* | rs2293348<br>rs4947492 | NA<br>NA | 1.9 × 10-2<br>5 × 10-2 |
| | Jandu et al. 2023 (102) | *Chr 5: 5q22.3* | rs75912034 | 1.24 | 1.21 × 10-10 |
| | | *Near ANOS1* | rs188287402 | 1.14 | 2.80 × 10-8 |
| | | *Near GPC5* | rs145328458<br>rs61966612 | 1.23<br>1.23 | 1.06 × 10-9<br>1.23 × 10-9 |
| | | *Near LINC01779* | rs77311050 | 1.34 | 2.54 × 10-8 |
| | | *LOC124901047* | rs12657177 | 1.24 | 1.12 × 10-10 |
| | | *Near PAX7* | rs643644 | 1.4 | 3.54 × 10-8 |
| | | *Near SLC10A4* | rs34063419 | 1.71 | 1.21 × 10-8 |
| | Kwok et al. 2022 (54) | *CYP2D6* | rs16947 | 3.82 | 4 × 10-2 |
| | Li et al. 2021 (99) | *ERCC5* | rs4771436<br>rs11069498<br>rs4150330 | 0.41<br>3.37<br>2.93 | 2.7 × 10-2<br>3 × 10-3<br>6 × 10-3 |
| | Liu et al. 2022 (107) | *SLC28A3* | rs885004 | 2.03 | 1 × 10-2 |
| | Meijer et al. 2021 (109) | *TCERG1L* | rs893507 | 3.11 | 5.3 × 10-10 |
| | Naderi et al. 2023 (108) | *DPPA4* | rs142667902 | NA | 1.7 × 10-7 |
| | Qi et al. 2022 (100) | *VEGF* | rs1570360 | HR 3.16 | 1.5 × 10-2 |
| | Ramos-Esquivel et al. 2020 (70) | *MTHFR* | rs1801133 | 1.69<br>1.56 | 5 × 10-3<br>1.3 × 10-2 |
| | Sharma et al. 2022 (57) | *ABCB1* | rs1128503<br>rs2032582 | 0.35<br>0.37 | 1 × 10-2<br>8 × 10-3 |

| | | | | | |
|---|---|---|---|---|---|
| | | *ABCC1* | rs41410450 | 0.2 | 4 × 10-2 |
| | Velasco-Ruiz et al. 2021 (106) | *POLMRT* | rs62134260 | 5.76 | 2.23 × 10-5 |
| | Wang et al. 2024 (94) | *SULT2A1* | rs182420 | 4.33 | 4.2 × 10-2 |
| | Yang and Liu 2020 (77) | *XPD* | rs13181 | 1.680 | 3.5 × 10-2 |
| | | *XRCC1* | rs25487 | 0.650 | 3.5 × 10-2 |
| | | *XRCC2* | rs3218556 | 0.605 | 1.9 × 10-2 |

Notes: HR, Hazard Ratio; NA, Not Available
