## Supplementary material for "From Genes to Personalized Cancer Care: A Systematic Review of Toxicity-Associated Genetic Variants in Solid Tumor Treatments": igure S1. Protective (OR<0.3) (A) and B) risky (OR>6) (B) SNVs forest plots

**Figure S1.** Protective (OR<0.3) (A) and B) risky (OR>6) (B) SNVs forest plots. Toxicities include irAEs (1), dermatological (2), dysphagia (3), gastrointestinal (4), hematological (5), leukopenia (6), neutropenia (7), thrombocytopenia (8), neuropathy (9) other toxicities (10).

A)

B)

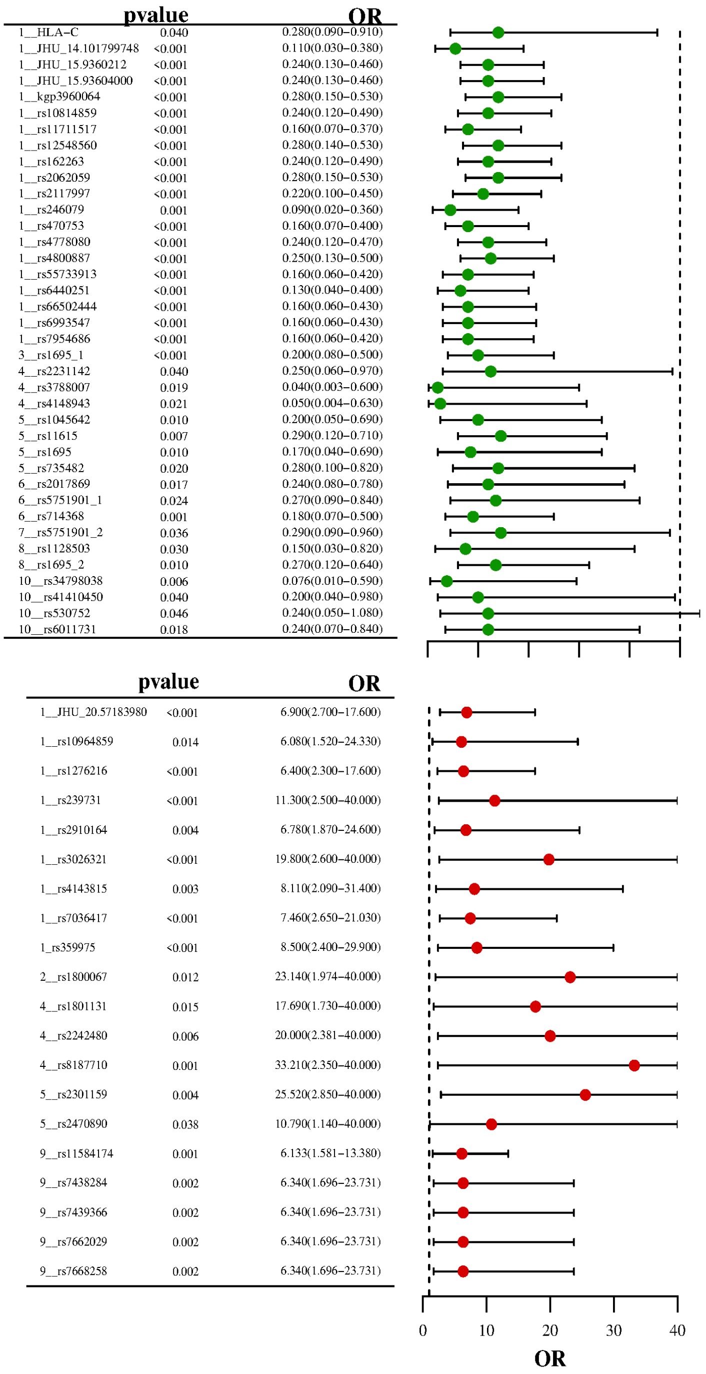

Supplement: igure S1. Protective (OR<0.3) (A) and B) risky (OR>6) (B) SNVs forest plots [file 338446_file06.pdf]
